# COVID-19: how the pandemic impacted the emergency department and urgent surgical activity in a medium-size Italian hospital

**DOI:** 10.1101/2020.11.19.20234856

**Authors:** Enrico Pinotti, Francesca Carissimi, Gianluca Baronio, Mauro Montuori, Deborah Ongaro, Michele Ciocca Vasino

## Abstract

**Backgrounds:** COVID-19 has grown rapidly in Lombardy, particularly in the province of Bergamo. To deal with the pressure the pandemic has exerted on the Italian health system; many hospitals have had to reorganize their medical and surgical activities. The aim of this study was to evaluate how the pandemic influenced the emergency department and urgent surgical activity in a medium-size hospital in the province of Bergamo.

**Methods:** In this retrospective observational study, we analyzed the number of admissions to the medical and surgical Emergency Room and their severity compared with those in the same period in previous years (2011-2019). Admission in the medical and surgical department and urgent surgical operation was also assessed.

**Results:** From March 7th to April 5th, 2020, we observe a reduction in emergency department access (−53%) when compared with the corresponding period of previous years. The number of medical admissions was similar to the past years (+0.9%), we observed a drastic reduction of surgical patients (−82.5%). We experienced a significant increase in hospitalizations in the medical department (+359%) and a reduction of admission in the surgical department (−71.2%).

**Conclusion:** SARS-CoV2 disease has spread so suddenly and severely that it has stressed Italian health system, in particular the Lombard one. Our data show the rise of critical medical ER accesses and the significant expansion in hospitalisation in the medical department with the necessary hospital reorganisation to face COVID-19 emergency. We also observed a reduction in both surgical ER accesses and urgent surgical activity.

## INTRODUCTION

The new severe acute respiratory syndrome coronavirus 2 (SARS-CoV-2 or COVID-19) originated in December 2019 in Wuhan, China ^1^, and then spread rapidly worldwide, until COVID-19 was WHO declared a pandemic on March 11th, 2020 ^2^. In Italy, Lombardy was the most affected region, the first case was identified on February 20th, 2020 ^3–5^. The emergency of the situation was clear from the beginning of virus spreading, so the government delineated red-zone areas where the citizens were isolated for 14 days to limit contact to try reduce the SARS-CoV-2 infection ^6^. Despite these measures, COVID-19 has grown following an exponential trend, so the Government declared a national lockdown in the night between 9th and 10th March 2020 ^7^. The movement of individuals in the whole Italian national territory was limited unless strictly motivated by work or health reasons. Schools, museums, cinemas, theatres, and any other social, recreational, or cultural centre must stay closed. Any gathering in public spaces is forbidden, including sporting events, church and funerals. Most shops must stay closed. Those selling essentials, such as supermarkets or pharmacies, need to ensure a distance of at least 1 m between customers ^8–10^. The province of Bergamo is located in Lombardy and has 1.12 million inhabitants, it was one of the most affected areas in Italy^11–13^ with 997 COVID-19 positive patients on 7th March and 9.815 on April 5th (0.8% of Bergamo population, Italian Ministry of Health). It should additionally be noted that the pharyngeal swab test was performed only on symptomatic patients, so the number of cases is probably more significant. The epidemiological analysis of Flaxman et al. estimated that in Italy cumulatively 5.9 million people have been infected as of March 28th, giving an attack rate of 9.8% of the population ^14^.

Policlinico San Pietro represents a hospital with 300 beds, located in the province of Bergamo. It was one of the firsts Italian hospitals transforming itself into a COVID-19 hospital. From March 7th to April 5th, 509 patients were hospitalized in the medicine department, almost all for bilateral interstitial pneumonia due to suspicion or confirmed COVID-19. To manage this number of patients, medical and intensive-care beds have been implemented, suspending all non-life-saving clinical activities. In our institute the elective surgical activity was suspended after the session of March 6th due to COVID-19 emergency; only urgent-emerging and life-saving surgery was active. The aim of this study was to evaluate how the emergency department and urgent surgical activity changed during the COVID-19 pandemic first month in a medium-sized hospital in a critical area.

## MATERIAL AND METHODS

In our institute the elective surgical activity was suspended after the session of March 6th due to COVID-19 emergency, we considered the period between March 7th and April 5th. In our emergency department, 30,537 people are evaluated every year (mean number of access in 2011-2019), about 84 patients per day. In our emergency department, patients are divided in two groups (medical and surgical patients) according to the suspected diagnosis at the entrance. If the suspected diagnosis is internalistic, cardiological or neurological, the patient is referred to the medical emergency room (ER) while if the suspected diagnosis refers to general surgery, orthopedics, traumatology, urology, neurosurgery, otolaryngology or ophthalmology is directed to the surgical ER. Patients are first evaluated at the check-in desk (Triage) and a color is assigned in accordance with the level of priority, clinical conditions and vital signs: RED for life-threatening conditions, YELLOW for potentially critical conditions, GREEN for minor injuries or illnesses, or WHITE for non-urgent conditions. We analyzed data from the first 30 days from the beginning of the COVID-19 pandemic in our hospital (San Pietro Polyclinic, Bergamo, Italy). Two authors (EP and GB) independently searched for numbers of emergency department access and classification by color in the period between March 7th and April 5th. Data on the last 10 years (from 2011 to 2020) were analyzed. The hospitalization criteria and the criteria for surgery intervention adopted during COVID-19 emergency are the same as those used in previous years. The admissions in surgical and medical ward from the emergency department and the number of urgent surgical operations were also assessed. In the evaluation of urgent surgical operations, only patients from ER were considered; reoperations after elective surgery, trauma and deferred emergencies were excluded. Local Ethical Committees review of the protocol deemed that formal approval was not required owing to the retrospective, observational, and anonymous nature of this study. Statistical analysis was performed with SPSS software (Statistical Package for Social Science).

## RESULTS

From March 7th to April 5th, 2020, during COVID-19 Pandemic 1191 patients presented to our emergency department (mean 39.7 pts/days). We observe 53% access reduction when compared with the corresponding period of previous years (mean number of admission to emergency department was 2534, 84.4 pts/days). Figure 1a. If the number of medical admissions was similar to the mean of the past nine years (904 patients in 2020 vs 896 patients in the period of 2011-2019; 30.1 vs 29.9 pts/days), we observed a drastic reduction of surgical patients. From March 7th to April 5th, 2011-2019, the mean number of admission in surgical ER was 1638 vs only 287 of 2020 with 82.5% reduction (54.6 vs 9.6 pts/days). Figure 1b. When we analyze color-code assigned to patients at the check-in desk (Triage) in accordance with the level of priority, clinical conditions and vital signs we notice a reduction of white and green-code patients (respectively -80.6% and -72.9%) when compared with the mean of the previous period; the number of yellow-code patients was similar 579 vs 614 (−5.8%) and there was an increase of critical red-code patients 101 vs 22 (+359%), Table 1. In the 30 days analyzed in 2020, we experienced a reduction in minor accesses in medical ER (−100% white-codes, -38.3% green-codes) and an increase in severe cases (+43.2% yellow-codes, +405.3% red-codes) when compared with the corresponding period of 2011-2019 (table 1). In the surgical emergency room, we observed a reduction in all the codes excluding the red ones (−75% white-codes, -86,7% green-codes, -64.7% yellow-codes and +78.6% red-codes). Thirteen of the 287 patients evaluated in surgical ER needed hospitalization in the general surgery ward. The mean hospitalization from surgical ER in the same period of 2011-2019 was 45 patients with 71.2% reduction. Figure 1c. Despite we suffered a reduction in admission to the general surgery department, in the period between March 7th and April 5th, 2020, 4.5% of patients evaluated in the surgical ER compared to a mean of 2.77% in 2011-2019 same period were hospitalized in general surgery. We observed a significant increase in hospitalizations in the medical department (+359%) with 509 hospitalizations in 2020 vs an mean of 111 in previous years (figure 1d).

**TABLE 1:** Access to Emergency department, admission to medical and surgical department

|  |  |  | 2011 | 2012 | 2013 | 2014 | 2015 | 2016 | 2017 | 2018 | 2019 | Mean<br>2011-2019 | 2020 | Variation |
| --- | --- | --- | --- | --- | --- | --- | --- | --- | --- | --- | --- | --- | --- | --- |
| Number of access to Emergency Department | Overall | 30 days<br>Mean for day | 2797<br>93.2 | 2669<br>89.0 | 2408<br>80.3 | 2456<br>81.9 | 2445<br>81.5 | 2369<br>79.0 | 2544<br>84.8 | 2425<br>80.8 | 2693<br>89.9 | 2534<br>84.5 | 1191<br>39.7 | -53.0% |
|  | Medical | 30 days<br>Mean for day | 982<br>32.7 | 889<br>29.6 | 842<br>28.1 | 871<br>29.0 | 868<br>28.9 | 868<br>28.9 | 861<br>28.7 | 920<br>30.7 | 959<br>32.0 | 896<br>29.9 | 904<br>30.1 | +0.9% |
|  | Surgical | 30 days<br>Mean for day | 1815<br>60.5 | 1780<br>59.3 | 1566<br>52.2 | 1585<br>52.8 | 1577<br>52.6 | 1501<br>50.0 | 1683<br>56.1 | 1505<br>50.2 | 1734<br>57.8 | 1638<br>54.6 | 287<br>9.6 | -82.6% |
| Color code access to emergency department (overall) | White | 30 days<br>Mean for day | 45<br>1.5 | 33<br>1.1 | 26<br>0.9 | 35<br>1.2 | 26<br>0.9 | 55<br>1.8 | 34<br>1.1 | 48<br>1.6 | 70<br>2.3 | 41<br>1.4 | 8<br>0.3 | -80.6% |
|  | Green | 30 days<br>Mean for day | 2036<br>67.9 | 2003<br>66.8 | 1745<br>58.2 | 1875<br>62.5 | 1813<br>60.4 | 1758<br>58.6 | 1875<br>62.5 | 1709<br>57.0 | 1892<br>63.1 | 1856<br>61.9 | 503<br>16.8 | -72.9% |
|  | Yellow | 30 days<br>Mean for day | 705<br>23.5 | 611<br>20.4 | 621<br>20.4 | 533<br>17.7 | 580<br>19.3 | 543<br>18.1 | 614<br>20.5 | 625<br>20.8 | 698<br>23.2 | 614<br>20.5 | 579<br>19.3 | -5.8% |
|  | Red | 30 days<br>Mean for day | 11<br>0.4 | 22<br>0.7 | 16<br>0.5 | 13<br>0.4 | 26<br>0.9 | 13<br>0.4 | 21<br>0.7 | 43<br>1.4 | 33<br>1.1 | 22<br>0.7 | 101<br>3.4 | +359% |
| Color code access to MEDICAL emergency room | White | 30 days<br>Mean for day | 12<br>0.4 | 11<br>0.3 | 6<br>0.2 | 6<br>0.2 | 4<br>0.1 | 7<br>0.2 | 10<br>0.3 | 13<br>0.4 | 15<br>0.5 | 9.3<br>0.3 | 0<br>0.0 | -100% |
|  | Green | 30 days<br>Mean for day | 534<br>17.8 | 533<br>17.8 | 490<br>16.3 | 533<br>17.8 | 517<br>17.2 | 539<br>18.0 | 520<br>17.3 | 531<br>17.7 | 575<br>19.2 | 530<br>17.7 | 327<br>10.9 | -38.3% |
|  | Yellow | 30 days<br>Mean for day | 425<br>14.2 | 324<br>10.8 | 333<br>11.1 | 319<br>10.6 | 324<br>10.8 | 310<br>10.3 | 315<br>10.5 | 339<br>11.3 | 343<br>11.4 | 336<br>11.2 | 481<br>16.0 | +43.2% |
|  | Red | 30 days<br>Mean for day | 11<br>0.37 | 21<br>0.70 | 13<br>0.43 | 13<br>0.43 | 23<br>0.77 | 12<br>0.40 | 16<br>0.53 | 37<br>1.23 | 26<br>0.87 | 19<br>0.64 | 96<br>3.2 | +405.3% |
| Color code access to SURGICAL emergency room | White | 30 days<br>Mean for day | 33<br>1.1 | 22<br>0.7 | 20<br>0.6 | 29<br>1.0 | 22<br>0.7 | 48<br>1.6 | 24<br>0.8 | 35<br>1.2 | 55<br>1.8 | 32<br>1.1 | 8<br>0.3 | -75% |
|  | Green | 30 days<br>Mean for day | 1502<br>50.1 | 1470<br>49.0 | 1255<br>41.8 | 1342<br>44.7 | 1296<br>43.2 | 1219<br>40.6 | 1355<br>45.1 | 1178<br>39.3 | 1317<br>43.9 | 1326<br>44.2 | 176<br>5.9 | -86.7% |
|  | Yellow | 30 days<br>Mean for day | 280<br>9.3 | 287<br>9.6 | 288<br>9.6 | 214<br>7.1 | 256<br>8.5 | 233<br>7.8 | 299<br>10.0 | 286<br>9.6 | 355<br>11.8 | 278<br>9.3 | 98<br>3.3 | -64.7% |
|  | Red | 30 days<br>Mean for day | 0<br>0.00 | 1<br>0.03 | 3<br>0.10 | 0<br>0.00 | 3<br>0.10 | 1<br>0.03 | 5<br>0.17 | 6<br>0.20 | 7<br>0.23 | 2.8<br>0.10 | 5<br>0.17 | +78.6% |
| Admission to Department of General Surgery |  | 30 days<br>Mean for day | 48<br>1.6 | 54<br>1.8 | 45<br>1.5 | 45<br>1.5 | 50<br>1.7 | 48<br>1.6 | 34<br>1.1 | 47<br>1.6 | 36<br>1.2 | 45<br>1.5 | 13<br>0.4 | -71.2% |
| Admission to Department of Medicine |  | 30 days<br>Mean for day | 86<br>2.9 | 105<br>3.5 | 95<br>3.2 | 119<br>4.0 | 118<br>3.9 | 131<br>4.4 | 119<br>4.0 | 106<br>3.5 | 122<br>4.1 | 111<br>3.7 | 509<br>17.0 | +359% |

**FIGURE 1:**
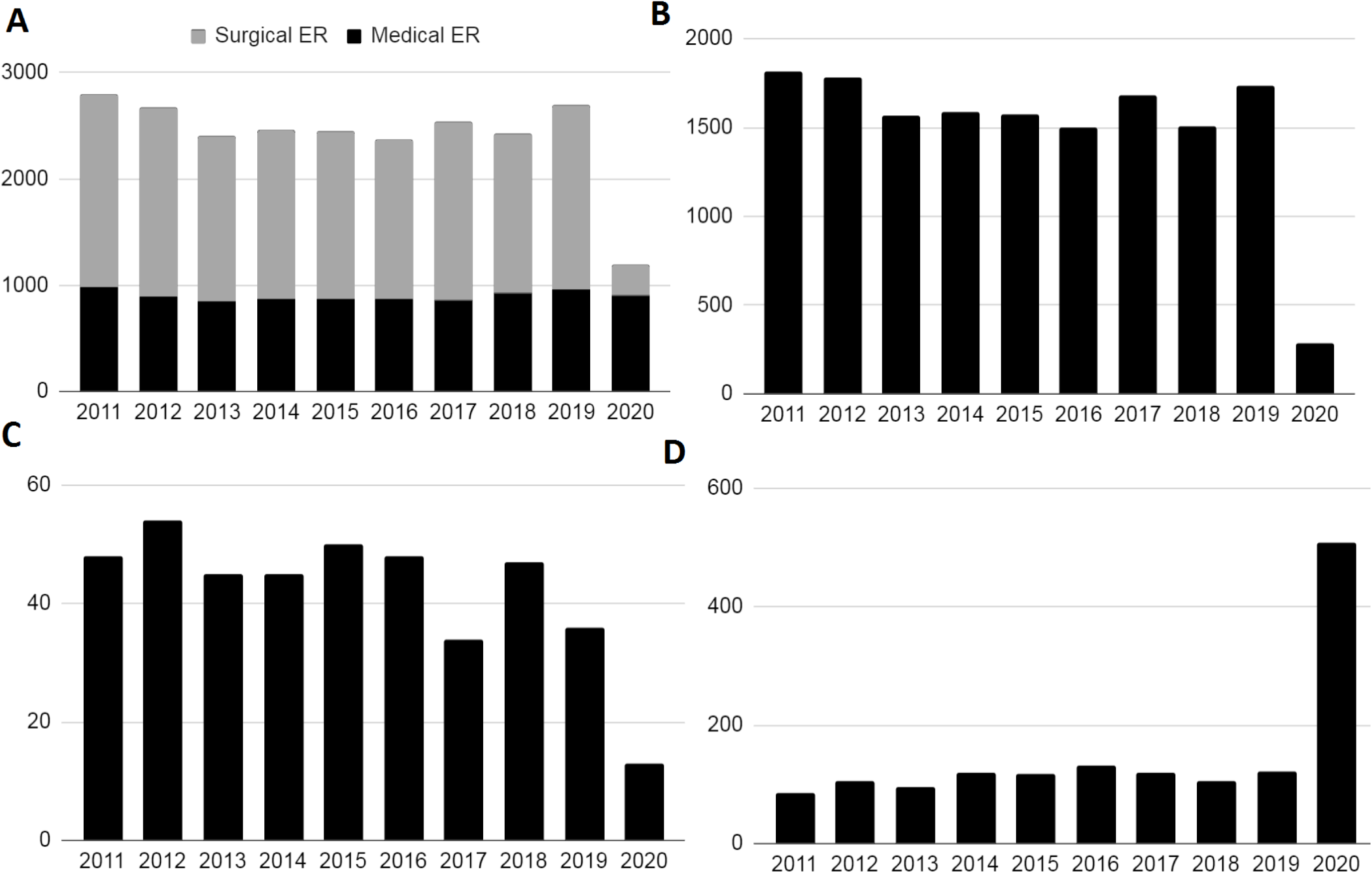
Figure 1a: Emergency room (medical and surgical) total admission Figure 1b: Emergency room (Surgical admission) Figure 1c: Admission to the general surgery ward Figure 1d: Admission to the medical ward

From March 7th to April 5th, 2020, we perform six urgent surgical operations: one patient underwent cholecystectomy for acute cholecystitis, one appendicectomy was performed for perforated appendicitis, two patients underwent hernia repair for incarcerated hernia and two colonic resections were performed (one for colonic ischemia and the other for diverticular perforation) (Table 2). When we analyze urgent-emergent surgical operations from March 7th to April 5th, 2011-2019, we exclude reoperation after elective surgery, trauma and deferred emergencies and consider only patients from ER. We notice a 47.5% reduction of urgent-emergent surgery when compared 2020 with the period from 2011 to 2019. For pathologies where conservative antibiotic treatment is possible (cholecystitis and appendicitis) the reduction was 60% while for the other pathologies the reduction was 37.9%.

**TABLE 2:** Urgent surgical operations

Urgent surgical operations
|  | 2011 | 2012 | 2013 | 2014 | 2015 | 2016 | 2017 | 2018 | 2019 | Mean<br>2011-2019 | 2020 | Variation |
| --- | --- | --- | --- | --- | --- | --- | --- | --- | --- | --- | --- | --- |
|  | 16 | 11 | 13 | 13 | 11 | 11 | 7 | 9 | 12 | 11.4 | 6 | -47.5% |
| Appendectomy | 6 | 2 | 2 | 3 | 5 | 3 | 3 | 2 | 4 | 3.3 | 1 | -70% |
| Cholecystectomy | 2 | 3 | 1 | 1 | 2 | 3 | 0 | 2 | 1 | 1.7 | 1 | -40% |
| Hernia repair | 1 | 4 | 0 | 3 | 0 | 2 | 1 | 2 | 3 | 1.8 | 2 | +12.5% |
| Colonic resection | 6 | 2 | 5 | 4 | 2 | 1 | 2 | 2 | 2 | 2.9 | 2 | -30.1% |
| Other | 1 | 0 | 5 | 2 | 2 | 2 | 1 | 1 | 2 | 1.8 | 0 | -100% |
| Pathologies with possible conservative treatment (appendicitis and cholecystitis) | 8 | 5 | 3 | 4 | 7 | 6 | 3 | 4 | 4 | 5.0 | 2 | -60.0% |
| Pathologies without possible conservative treatment (others) | 8 | 6 | 10 | 9 | 4 | 5 | 4 | 5 | 8 | 6.4 | 4 | -37.9% |

## DISCUSSION

SARS-CoV2 disease has spread so suddenly and severely that it has stressed Italian health system, in particular the Lombard one, that had taken extraordinary measures to address the constant patients increase in medical departments ^15^, as our data show in table 1. These measures took place on two fronts: protection of healthcare personnel to avoid intra-hospital COVID-19 diffusion (supply of Personal Protective Equipment, body temperature check before each work shift, nasal swab in case of symptoms even mild) and creation of COVID-dedicated beds. All departments blocked the elective activity to be able to meet the beds demand from medical ER, which in our center were about 3.5-fold greater than in the same period in the years 2011-2019. In particular, in the general surgery department most patients, when clinically feasible, were discharged; all the elective surgery was suspended, and the operating rooms were transformed into intensive-care beds; the number of attending surgeons on the ward has reduced to cover work shifts in the emergency room and medical department. Only 1 on 6 operating rooms was maintained for eventual emergent/urgent surgery. Most of the Lombardy hospitals were mainly dedicated to coronavirus patients. The Lombardy Region has identified 18 HUBs, hospitals reserved for major trauma, heart attacks, strokes and other emergencies. All other hospitals, including ours have been classified as SPOKE intended for the treatment of COVID-19 patients, especially intensive-care units. Therefore, we decided not to include in our population the patients that in the period between 2011-2019 had been classified as trauma, to minimize the selection bias. During the 30 days taken into consideration, there have been deep changes in ER access, we have seen a reduction in minor codes, white and green, by 80.6% and 72.9% respectively, with a substantial stability of the yellow codes (−5.8%) and red codes increase of 359%. Even more interesting was the evaluation of ER access investigating medical and surgical area separately. Surgical ER underwent to reduction of approximately 83% in the total amount of accesses, in detail -75%, -86.7 and -64.7% respectively for white, green and yellow codes; on the other hand, the data on red codes seem to go against the trend, showing +78.6% raise; obviously the number is small to have an effective estimate of the real increase. This access codes redistribution could be explained by general practitioners trying conservative treatment at home and patients’ more expected attitude for fear of infection in overcrowded ER. During the 30 days in the study, medical ER was deeply stressed (table 1), not for an overall admission increase that was substantially stable (+0.9%), indeed we observed a drastic reduction for minimal injuries patients (−100% white codes, -38.3% green codes), a notable increase of significant clinical conditions (+43.2% yellow codes) and a dramatic expansion of patients with life-threatening conditions (+405.3% red codes). If the Health Minister ordinance to stay at home and contact general practitioners if flu symptoms and fever (up to 37.5°C) are present to avoid ER overcrowding, could explain the reduction of minor codes; the severity and rapid onset of COVID-19 symptoms may justify the raise of critical conditions. Of course, this change in the type of access (medical vs surgical) and in the distribution of codes with a shift to the major codes leads accordingly to a greater number of admissions in medical ward, approximately 360% increase (table 1); to meet this demand, the departments suspended their elective activities; on the other hand, due to reduced access to surgical ER, admissions to emergency surgery decreased by 71.2%, indeed the reduction in surgical activity was much greater, considering also elective activity suspension. In this scenario, the impact was also on emergency surgeries that overall had a drop of 47.5% (table 2); however, there is a significant difference in the reduction of interventions depending on pathologies taken into consideration; in fact, for pathologies where a conservative approach is not viable, the surgery number has remained almost stable, while pathologies with possible conservative treatment have 60% decrease. In this dramatic situation our approach has been to attempt conservative treatment where possible and to reschedule the subsequent surgery in a elective regime to avoid prolonged hospitalization with relative infectious consequences. Appendicitis and cholecystitis in which conservative treatment is possible, the surgery has decreased by 70% and 40% respectively; for these illnesses often due to an infection, according to guidelines ^16^, conservative treatment with antibiotic therapy is safe and feasible, even at home in selected cases; for cholecystitis, another therapeutic option is cholecystostomy, which has proven to be a valid alternative to cholecystectomy in fragile patients who are not responsive to the antibiotic ^17^. Incarcerated/strangulated hernia and bowel obstruction/perforation don’t seem to have reduction or increase, unfortunately population is to small to perform reliable statistics analysis, because these conditions cause a a life-threatening condition that needs emergency surgery, in fact conservative treatment is often not conclusive for these patients ^18–20^. When surgery could not be postponed, measures were taken during hospital stay to avoid any intra-hospital infection even in patients without respiratory symptoms ^21,22^: hospitalization in covid free wards, patients’ use of a mask, isolation in the room, healthcare personnel Personal Protective Equipment use.

## CONCLUSION

The result of COVID-19 pandemic on Italian healthcare has been dramatic, we analyzed the first 30-days of pandemic in a medium-size Lombard hospital. Our results show the rise of critical medical ER accesses and the significant expansion in hospitalisation in the medical department with the necessary hospital reorganisation to face COVID-19 emergency. We also observed a reduction in both surgical ER accesses and urgent surgical activity, with conservative management as the first option where feasible. However, the number of pathologies where an emergency surgical approach is necessary has not decreased significantly compared to previous years. In the first 30-days of COVID-19 pandemic, despite the overall ER access was stable when compared with previous years, it was stressed due to an significantly increase of patients with critical and life-threatening conditions associated with a drastic minor-code patients reduction.

## Data Availability

All data are avable

## ACKNOWLEDGMENTS

None

